# How to make calibration less painful – a proposition of an automatic, reliable and time-efficient procedure

**DOI:** 10.1101/2022.10.03.22280662

**Authors:** Karolina Swider, Ricardo Bruña, Stephan Moratti

## Abstract

**Background:** In neurophysiological pain studies, multiple types of calibration methods are used to quantify the individual pain sensation stimuli that have different modalities. However, such studies often lack calibration procedure implementation, have a vague protocol description, do not provide data quality quantification, or even omit required control for gender pain differences. All this hampers not only study repetition but also interexperimental comparisons. Moreover, typical calibration procedures are long and require a high number of stimulations which may cause participants’ discomfort and stimuli habituation.

**Method:** To overcome those shortcomings, we present an automatic staircase pain calibration method for A-delta-specific electrical stimulation adjusted to the magnetoencephalography environment. We provide an in-depth data analysis of the collected self-reports from seventy healthy volunteers (37 males) and propose a method based on a dynamic truncated linear regression model (tLRM). We compare its estimates for the sensation (*t*), and pain (*T*) thresholds, as well as for the mid-pain stimulation (*MP*), with those calculated using a traditional threshold method and standard linear regression models.

**Results:** Compared to the other threshold methods, tLRM exhibits higher R^2^ and requires 36% fewer stimuli application and has significantly higher *t* and lower *T* and *MP* intensities. Regarding sex differences, both lower *t* and *T* were found for females compared to males, regardless of the calibration method.

**Conclusions:** The proposed tLRM method quantifies the quality of the calibration procedure, minimizes its duration and invasiveness, as well as provides validation of linearity between stimuli intensity and subjective scores, making it an enabling technique for further studies. Moreover, our results highlight the importance of control for gender in pain studies.

**Summary:** The purpose of this study was to shorten and automatize the calibration method which is an enabling technique for realizing neurophysiological studies on pain. The proposed method is based on a dynamic truncated linear regression model and was shown to require 36% fewer stimuli application compared to the traditional staircase method. Furthermore, the calibration was adjusted to A-delta specific intraepidermal electrical stimulation, quantifies the quality of the resulting calibration parameters and provides a validation of linearity between stimuli intensity and subjective scores. The results also highlight the importance of control for participant gender in studies where different types of stimulation are used to induce pain sensation.

## 1. Introduction

Pain perception is a subjective experience, influenced by psychosocial, affective, cognitive, and biological factors^1,2^. To investigate pain processing and perception, standardized stimuli of different modalities (e.g. mechanical, electrical, thermal) are used to produce controlled pain sensation^3^. Essentially, such studies require a calibration procedure able to map the objective stimulation intensity to its subjective individual perception (e.g., the sensation *t*, and pain, *T*, thresholds).

The calibration protocols^4^, as well as modalities and characteristics of pain stimulation, highly differ from one study to another^3^, making comparisons between them challenging. A well-designed calibration procedure should be characterized by three factors: standardization (unified calibration protocols for specific stimulus types), automatization (environmental factors control and high replicability), and detailed description of its implementation (e.g. duration, stimuli number, rejection criteria). Unfortunately, this information is rarely provided, and the discussion of data is often neglected or, in the best case, scarce. An additional problem is the nature of pain assessment. Despite of attempts to find better suited methods^5–8^, in both laboratory and clinical practice, subjective one-dimensional scales are used, which are known to suffer from certain limitations and weaknesses^9^. For instance, there are serious concerns about the linearity of the pain scores, hampering the use of mathematical calculations^10^ to extrapolate values out of the calibration range, or otherwise forcing translations of complex non-linear perception onto an inaccurate linear scale^11^. The reach of these limitations is inconclusive, as some studies show that numerical rating scales (NRS) behave as a ratio scales^12,13^, while other works deny it^14,15^. Moreover, the results of the linearity tests are reported only in a few studies^14,16,17^.

Consequently, calibration can be considered as one of the most demanding and important parts of pain research. In this work, we propose a novel, completely automatic calibration method, accompanied with metrics to quantifying its accuracy and reliability, which are seldomly reported in pain studies. We compare the values of *T* and *t*, both computed via classical approaches, as the threshold-based method (TM) and via a family of linear regression methods (LRM), with the values obtained using our approach, with the objective of identifying the method that minimizes the number of painful stimuli applied in the calibration stage. Our proposal is based on LRM but, instead of using all the data from the calibration phase, it is a truncated LRM (tLRM) which analyses data up to an optimal point, chosen according to the maximum goodness-of-fit (R^2^) of the predictive model. Optimal R^2^, together with a convergence parameter based on the regression line gradient, formed the basis for rejection criteria, that is, the determination of a failed procedure.

tLRM aims to minimalize stimuli number and intensity to decrease participants’ discomfort during the calibration and helps avoiding habituation to painful stimuli, negatively impacting the results of the experimental phase^18^. Moreover, it ensures a higher degree of linearity, as habituation may introduce non-linearities to the NRS ratings, usually resulting in an overestimation of the intensity of the *T* threshold. Based on this, we predict that values of *T* calculated using TM and LRM will be higher than those calculated using the proposed tLRM. Furthermore, in laboratory studies it is common to define the stimulation intensity using a linear function of *T*^19,20^ even extrapolating scores higher than *T*^21,22^. We aimed to present and discuss a comparison of mid-painful stimulation calculated using our linear regression family.

Finally, although it is known that there are sex differences in pain sensitivity, their direction and scope are not always consistent across experimental pain modalities^23,24^. We aimed to evaluate sex differences in both *t* and *T* values for relatively new Adela-specific intraepidermal electrical stimulation (IES)^25^ what, to the best of our knowledge, has not been done before.

## 2. Methods

### 2.1. Participants

Seventy healthy volunteers (37 males; age mean±SD: 25.47±5.34) participated in the calibration phase of a magnetoencephalography (MEG) study, that will be reported elsewhere. The sample size was calculated prior to the MEG study using a GPower 3.1.9.4 based on the previous pain ^26,27^ and MEG ^28^. The optimal total sample size was shown to be between 42 and However, due to the possibility of participant rejection (for example, in electroencephalography (EEG) studies the rejection percentage is 28% ^29^) and the necessity of additional stimuli counterbalancing, this number was increased to 60. The extra 10 measurements were collected during pilot study. During data processing, four participants were excluded (see *Data analysis* below), resulting in 66 individuals (35 males; age between 18 and 36, mean±SD: 25.62±5.34).

Participants were recruited using the project website (https://neuroconmsca.wordpress.com/), and consisted of students and staff of the Complutense University of Madrid (UCM), the Centre for Biomedical Technology in Madrid (CTB), and the Basque Center on Cognition, Brain and language (BCBL) in San Sebastian. With respect to the MEG main experiment there were specific exclusion criteria which include: age below 18 and above 36 years, pregnancy, chronic diseases including chronic pain or migraine, recurrent pain, neurological or psychiatric diseases, heart disease, repeated unconsciousness, external and internal tissue damage, use of any type of medication or drug (psychoactive medication/substances such as antidepressants, antiepileptics, antipsychotics or illegal drugs), family history of epilepsy/photic epilepsy episode, claustrophobia, left-handedness, implantation of metal elements (e.g. endoprostheses, implants, metallic staples) and active implants (e.g.: pacemaker, neurostimulator, insulin pump, ossicle prosthesis), metal wire behind teeth and tattoos. Additionally, only participants who scored less than 8 on the *Hospital Anxiety and Depression Scale* (HADS) ^30^ and did not undergo a magnetic resonance study for 48 hours before the experiment could sign up to the study. All participants were encouraged to ask questions, were informed that they could resign form participation in any moment of the study and were guaranteed to receive 10 euros as compensation.

The calibration procedure design was approved by the Ethics Committee of the Universidad Complutense de Madrid (UCM), the Universidad Politécnica de Madrid (UPM), and the Basque Centre on Cognition, Brain and Language (BCBL), and followed the Helsinki Declaration and national and European Union regulations as part of a the MSCA project. None of the participants showed any signs of tissue damage nor reported genuinely adverse experiences as a result of undergoing the calibration procedure.

### 2.2. Stimulation hardware

The stimulation was controlled from a Debian PC running a MATLAB script with the use of Psychtoolbox library. Painful stimulation was controlled via the Elekta Stimulus Trigger Interface (STI102) StimBox, which is equipped with 16 binary input/output channels (BNC sockets) which generate a set of 16 5V analogue signals. We used two channels to control the stimulation strength. The complete stimulation hardware setup included: (1) PC with Windows 10 and LabVIEW, (2) NI myDAQ with Florida Research Instruments Inc. myDAQ BNC adapter for x10 oscilloscope probes with connectors, (3) Digitimer DS5 Bipolar Constant Current Stimulator, (4) WASP electrodes with connection cables, and (5) 3 BNC cables. The way all the hardware pieces were connected is shown in the Figure 1 and further described below.

**Figure 1.**
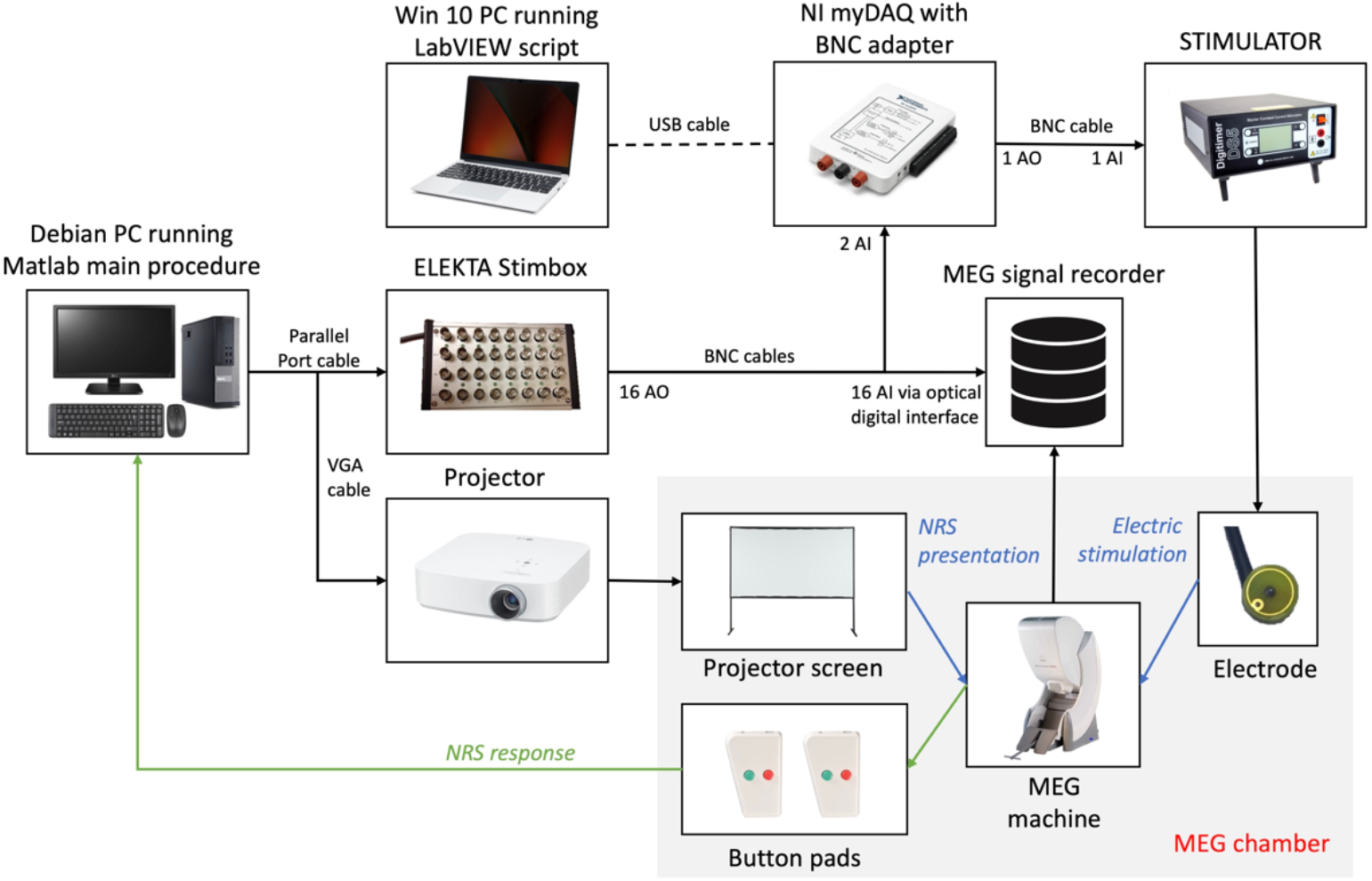
shows the main setup for the procedure. *Note*. Presented set up was used at CTB. Note that for the 24 participants that participated in the study at BCBL, Vipxx system (https://vpixx.com/) for stimulation was used instead of StimBox, and Windows computer was used instead of Debian 10 system.

We used WASP electrodes (Brainbox, United Kingdom), which selectively activate Adelta nociceptors, with very small stimulation intensity, approximately 0.01–2.0 mA^31^. By pressing the electrode, the needle tip (0.2 mm) is inserted into the skin, adjacent to the free nerve endings of the thin myelinated fibres in the epidermis and superficial part of the dermis^32,33^. The stimulus strength is set according to temporal summation of a long continuous duration^22^ or pulse train^25^ of fixed intensity. For this reason, WASP electrodes have a lower probability of generating adverse secondary effects like skin irritation. During calibration, two WASP electrodes were attached to the dorsum of the left hand^31^ with a spacing of 8 cm, but only one electrode was active and used to deliver 300 ms stimulations of ascending and descending intensities.

Digitimer DS5 Bipolar Constant Current Stimulator was used due to the possibility to control its output via analogue voltage input. The stimulator was placed outside of the MEG chamber to avoid electromagnetic interferences. It was driven by a NI myDAQ Data Acquisition Device (National Instruments, Austin, TX, USA) in combination with a myDAQ BNC adapter (Florida Research Instruments Inc., Cocoa Beach, FL, USA), as it accepted these TTL (transistor-to-transistor logic) signals of 5V as input and generated a varying voltage output. NI myDAQ was controlled via USB by a PC running a LabVIEW (National Instruments, Austin, TX, USA) script, which was running continuously, listening to the input channels at 1 kHz sampling rate and generating the requested output, if any (see Table S1 and Figure S1 in supplement materials).

### 2.3. Procedure

The participants were asked to enter the magnetically shielded room, take a seat in the MEG chair, facing a projector screen approximately 132 cm away, and have their hands placed on a board in front of them. They were informed that they would receive multiple increasing and decreasing intensities of electrical stimuli starting from 0mA, and that they would need to score their sensation according to an 11-point NRS using two 2-button response pads (Current Design, USA). In the meantime, the operator cleaned their skin with a cloth soaked in alcohol and attached the WASP electrodes to the dorsum of the left hand^31^. After that, the participants were left alone in the MEG chamber, but with permanent visual and verbal contact via a video system during all calibration procedure.

The procedure began with the NRS practice exercise where the participants learned how to use 2-Button Response Pads to choose the correct number on the NRS. First, they were provided with a written description of the exercise on the projector screen and then they were given 4 practice trials during which they needed to choose four numbers (e.g. 2, 5, 7, 4) on the scale. If they made a mistake, the trial was repeated. After the exercise, the participants underwent the calibration procedure which was completely automatic. The procedure applied electrical stimuli according to the timeline presented in Figure 2a, and followed the well-known staircase method, shown in detail in Figure 2b.

**Figure 2.**
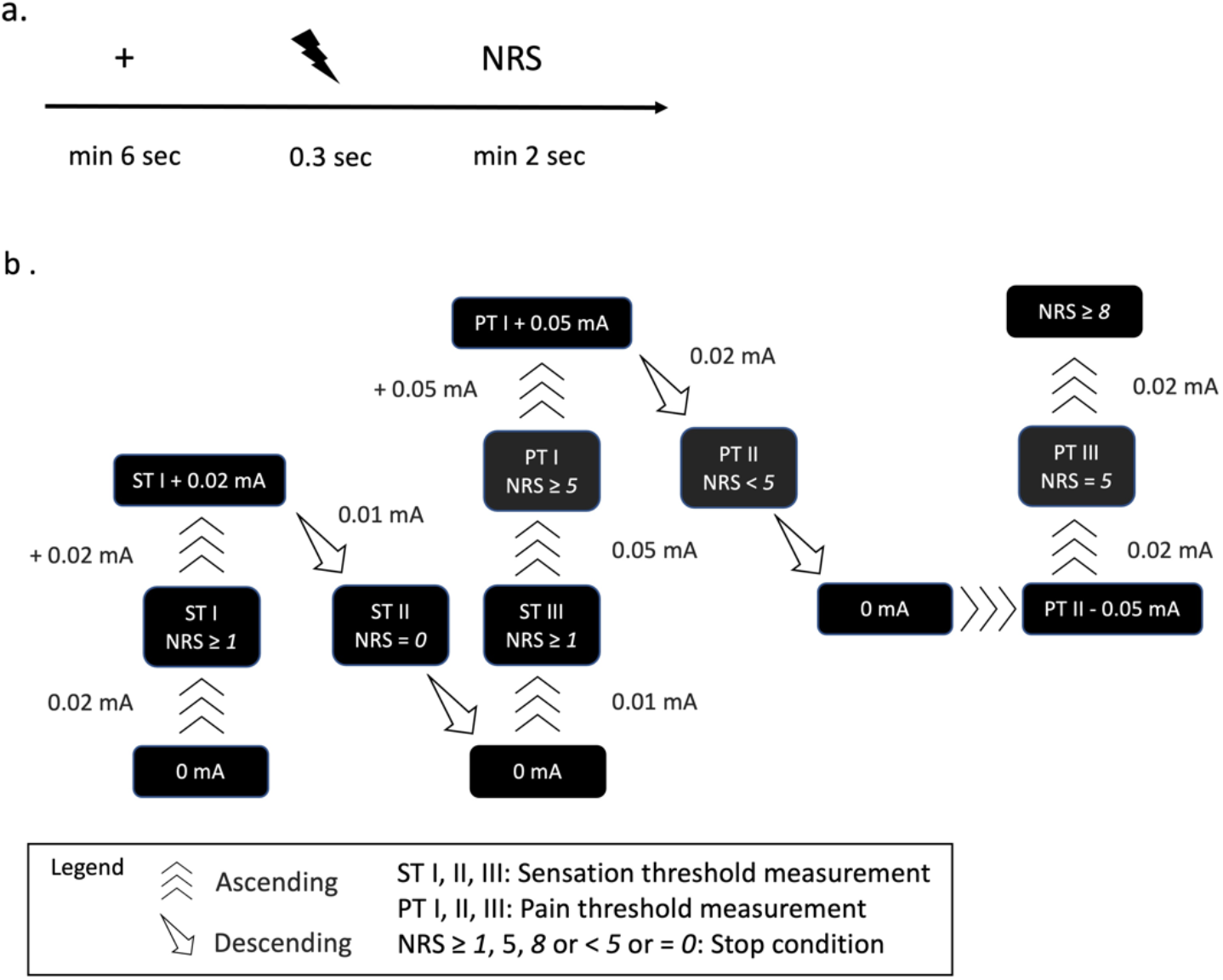
Calibration procedure. *Note*. a. Calibration timeline; b. Staircase calibration procedure, which used three increasing or decreasing intensities steps (small: 0.01 mA, medium: 0.02 mA and large 0.05 mA). Sensation (t) and pain threshold (T) intensities were established by staircase procedure, taking an average of the three readings (ST I, STII and STIII for t and PT I, PTII and PTIII for T) corresponding to NRS rating 1 and 5, respectively.

First, the participant was presented with a fixation cross for a minimum of 6 seconds. Then, the electric stimulation, with the current level of intensity, was applied, indicated in Figure 2a by a lightning bold icon. Thereafter, the participant was asked to rank the stimulus pain level, if any. After each stimulus, the participants were scoring their sensations on the 11-point NRS ranging from 0 – ‘*No electrical sensation’*, 1 – *‘I start to feel something’*, 5 *– ‘It starts to be painful’* to 10 – ‘*The strongest painful sensation imaginable’*, using the response pads. The numbers 1, 5 and 10 on the scale indicated the sensation threshold (t), the pain threshold (T), and the maximum pain tolerance, respectively. This scale resembles the one used in the other studies^21,34^.

Finally, the current level of intensity of the stimulation was increased or decreased, depending on the participant’s response. To reduce the probability of attenuation, and to reduce the number of steps required to achieve the t and T values, we used three different ascending/descending steps (0.01, 0.02, and 0.05, see Figure 2b). The interstimulus interval (ISI) was kept at minimum 8 sec, to further minimalize the probability of habituation.

Figure 3 presents an example of the output of the data collected during the calibration procedure.

**Figure 3.**
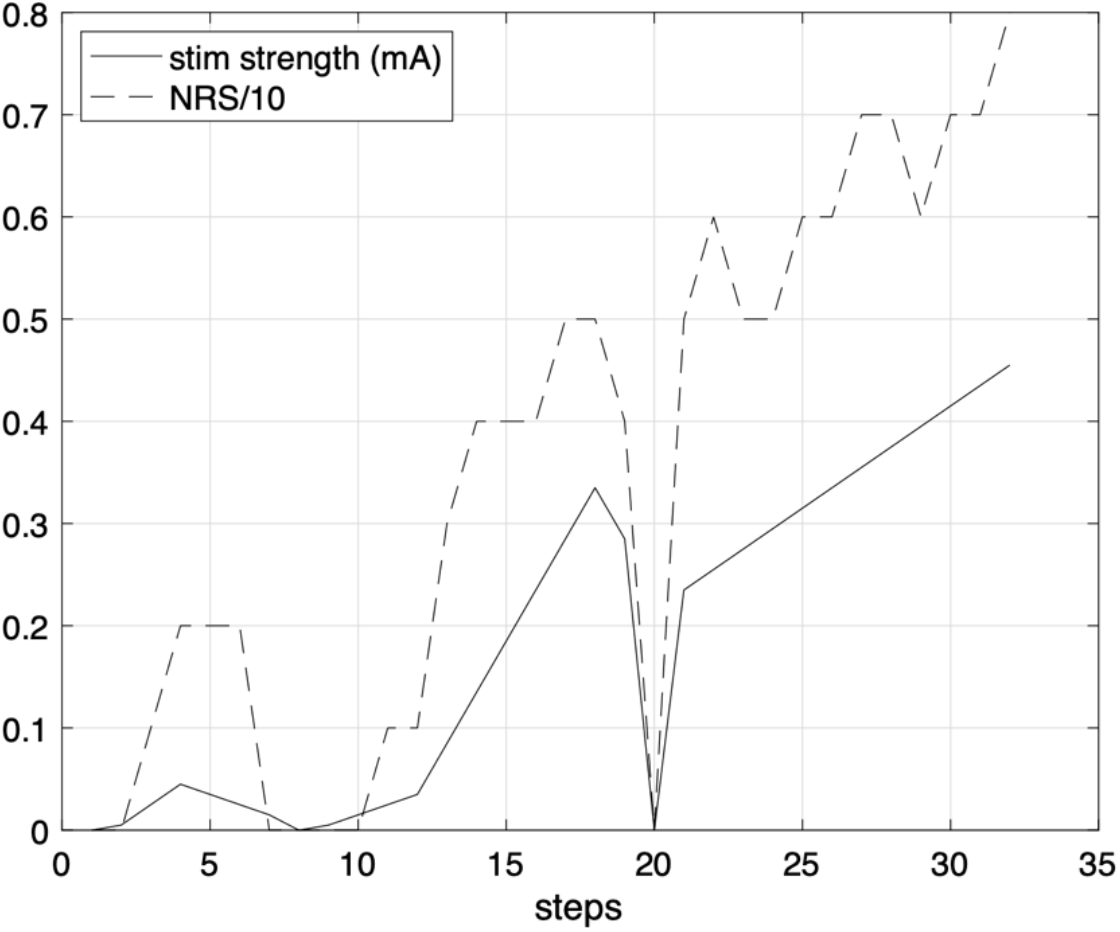
An example output provided at the end of the calibration procedure. *Note*. X-axis represents the number of steps and Y-axis is the simulation current in mA (black line) and the response NRS divided by 10 (black dashed line).

The graph was presented to the participant and served as a visual cue to check if the data exhibited a habituation pattern. The maximal stimulation that the participants received was that corresponding to an NRS of 8. In a post-test, 4 min after the completed calibration, to verify if the correct stimuli intensities were calculated for each participant, three intensities of different strength were delivered in ascending order. Participants were asked to verbally rate stimuli intensity using the same NRS scale. The stimuli intensities were calculated based on LRM and the following equation: 3*t* mA for tactile, 1.5*T*+0.02 mA for low-painful (LP) and LP*1.5 mA for mid-painful (MP).

### 2.4. Calibration methods

We calculated the *t* and *T* values using four methods: threshold method (TM), linear regression method (LRM) applied to the whole dataset, and two versions of the truncated linear regression method (tLRM), based on LRM.

#### 2.4.1. Threshold method (TM)

The threshold method calculates *t* by averaging three stimulation intensities (ST I, ST II, ST III) rated as 1 on the NRS (see Figure 2b), replicating a procedure used in previous studies^19,35^. ST I was achieved in the first ascending curve, selecting the first stimulation current with an NRS equal to 1. ST II was taken in the first descending curve, just before the first NRS rating equal to 0. ST III was achieved in the second ascending curve, when NRS reached 1.

Analogously, *T* was calculated by averaging three stimulation currents (PT I, PT II, PT III) scored as 5 on the NRS (see Figure 2b). PT I was achieved in the second ascending curve, selecting the stimulation current corresponding to an NRS equal to 5. PT II was taken in the second descending curve, just before an NRS equal to 4. PT III was achieved in the third ascending curve, selecting the stimulation current corresponding to an NRS equal to 5.

#### 2.4.2. Linear Regression Method (LRM)

We used a linear regression to model NRS ratings as a function of the stimulation intensity. We used the Matlab function *fitlm*, which fits a linear regression model to variables in the entire dataset and returns a relation *NRS = m·I+c*, where *I* is the stimulation current, *m* is the gradient, and *c* is the intercept. We also obtained *R*^*2*^ as a statistical measure to determine the proportion of variance in the dependent variable that can be explained by the independent variable.

#### 2.4.3. Truncated Linear Regression Method (tLRM)

In this work we propose two novel calibration methods, namely tLRMm and tLRMmc, obtained by modifying the LRM method to use only a specific part of the calibration data. The difference between both methods is that tLRMm assumes that the c intercept is zero, while tLRMmc does not. As previously, we used the Matlab function *fitlm* to create the regression models.

In both, the linear model is obtained not once, like in the case of LRM, but iteratively when a new data point is collected, starting from the second stimulation. As a result, *m, c*, and *R*^*2*^ are dynamically obtained as a vector of size *n-1*, where *n* is the total number of stimulations. Later, we truncate the vector based on the position of maximum *R*^*2*^, given that calibration was run for sufficiently many steps. To determine it, the *R*^*2*^ vectors for all the participants were plotted against the steps normalized with the second time a zero current stimulation was delivered. A moving average window of 250 data points was applied to interpret the data (see Figure 4).

**Figure 4.**
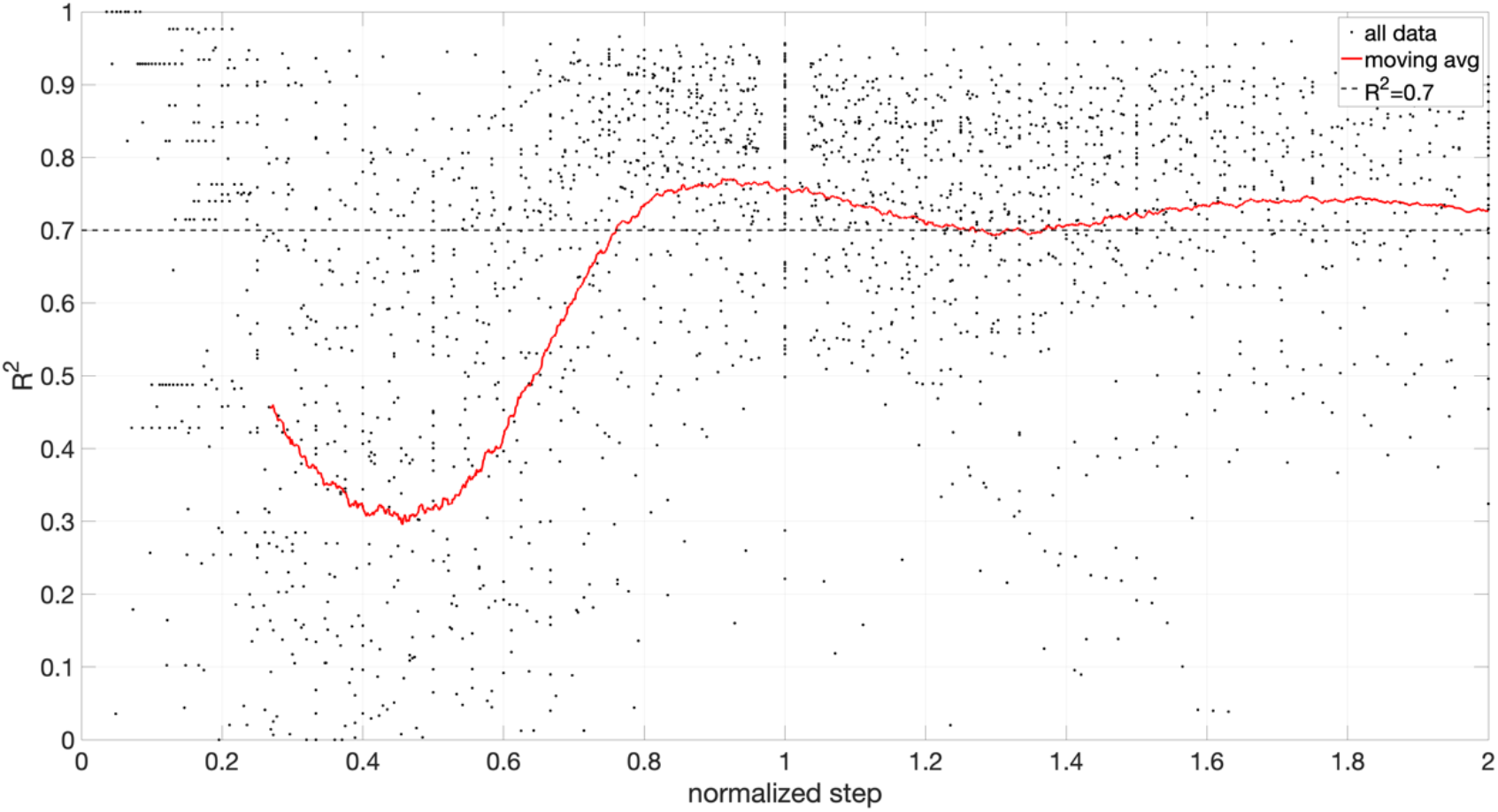
R^2^ values against number of steps normalized with the zero current simulation. *Note*. The black dots represent all the R^2^ values, the red line moving average over 250 points and dotted line the R^2^ =0.7.

According to Figure 4, the maximum *R*^*2*^ occurs at normalized step 0.91 and is equal to 0.77. For simplicity the second zero current stimulation, *i*, was chosen as the reference point, corresponding to normalized step 1 with *R*^*2*^ equal to 0.76. Additionally, to ensure optimal performance, a step range was defined between *i-3* and *i+3* and the optimal step for tLRM corresponded to the maximum *R*^*2*^ (see an example shown in Figure 5). Such design minimizes the number of painful stimuli and limits the highest pain sensation NRS rating to approximately 5, the pain threshold.

**Figure 5.**
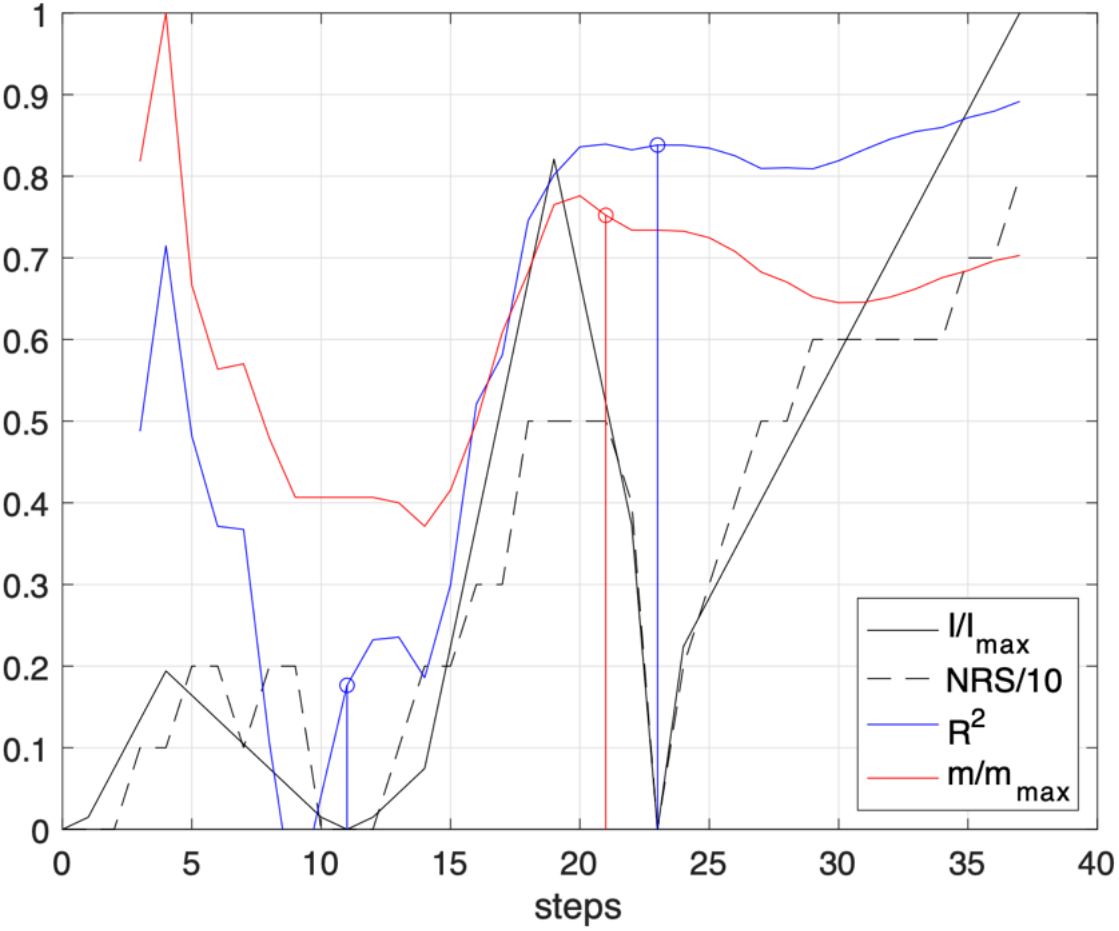
An example of tLRM output. *Note*. The x-axis represents calibration steps, starting from 0. The y-axis corresponds to: stimuli strength, I, normalized by its maximum value (black line); corresponding NRS rating divided by 10 (black dashed line); evolution of the coefficient R2 for the linear regression; and linear regression equation gradient, m, normalized by its maximum value (red line). Blue vertical lines represent points where zero current stimulation was applied, and the red vertical line represents the optimal truncation point.

## 3. Data analysis

### 3.1. Rejection criteria

To ensure that only good-quality data were analysed, we quantitatively checked which participants ‘failed’ the calibration procedure. This verification was performed for the tLRMmc and tLRMm, as the evaluation of these methods is the object of this study.

We propose three rejection criteria with their corresponding thresholds. The first one is based on *R*^*2*^ where high values indicate good data linearity. A recent study on thermal stimulation^36^ established that for a successful calibration procedure *R*^*2*^ should be at least 0.4. The second is related to the convergence of the parameters. If the calibration procedure is stopped at a point where the regression model parameters are invariant of collecting more data, we can say that the calibration procedure has converged. As a measure of convergence, we propose to modify the classical Cauchy’s criterion based on the gradient *m*:

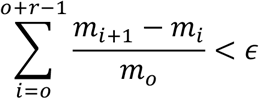

where *o* subscript symbolizes the optimal truncation step and ϵ is a limit which we propose to set to 0.25. Finally, the third criterium limits the stimulation current to prevent participants from receiving multiple pain stimulations of very high intensity and habituation). In our case, we chose an upper limit to the pain sensation (NRS=5) as 0.635 mA.

### 3.2. Statistical analysis

We investigated if there are differences in the goodness of fit for the three linear regression models (LRM, tLRMm and tLRMmc). *R*^*2*^ distributions were negatively skewed (-1.41, -1.24 and -1.31 respectively) and did not satisfy the Anderson-Darling test for normality (p<0.005). To mitigate the skewness, a set of 40 different power transformations was tested, which concluded that power 4.69 yielded optimal normalization results (skewness of 0.24, - 0.30, and -0.35, respectively and Anderson-Darling normality test p=0.054, 0.040, and 0.055, respectively). Next, we performed a mixed effects repeated measure ANOVA for R^2^ with *Linear Regression Type* (LRM, tLRMm, tLRMmc) as a within- and *Gender* as between-subject factor (females and males).

To indicate which calibration method requires less stimuli, we used a nonparametric Wilcoxon signed rank test on the stimuli number required by LRM vs tLRM (by definition, the tLRMm and tLRMmc require the same number of stimuli).

Afterward, we investigated if there were differences between the four tested calibration methods. We performed a mixed effects repeated measure ANOVA for the calculated values of *T* and *t* with *Threshold Type* (sensation and pain threshold) and *Gender* (females and males) as between- and *Calibration Type* (TM, LRM, tLRMm, tLRMmc,) and *Threshold Typ*e (t and T) as within-subject factors.

Next, we compared the intensity of mid-pain stimulation calculated for each of the calibration method. For TM it was obtained by *1.5*T* used in previous studies^20,26^. For linear models, the intensity leading to an NRS of 8 was used. Then, we performed a repeated measures ANOVA for mid-pain intensity (mA) with *Calibration Type* (TM, LRM, tLRMm, tLRMmc) as within- and *Gender* (females and males) as between-subject factor.

If necessary, Greenhouse-Geisser correction was applied for violations of sphericity, and in the case of interaction effects, the repeated measure ANOVA was followed by multiple comparison tests, using Bonferroni correction for multiple comparisons, to indicate the meaning of the effect. Data analysis was performed using MATLAB (R2020b Update 2) and statistical analysis were performed using IBM SPSS (Version 26).

## 4. Results

According to the rejection criteria, we determined that 4 participants (5.7%) need to be excluded from further analysis due to low *R*^*2*^ (below 0.4, see Figures 6a and 6d), stimulation currents above 0.635mA for NRS of 5 (see Figure 6b), or lack of convergence (see Figures 6c and 6d).

**Figure 6.**
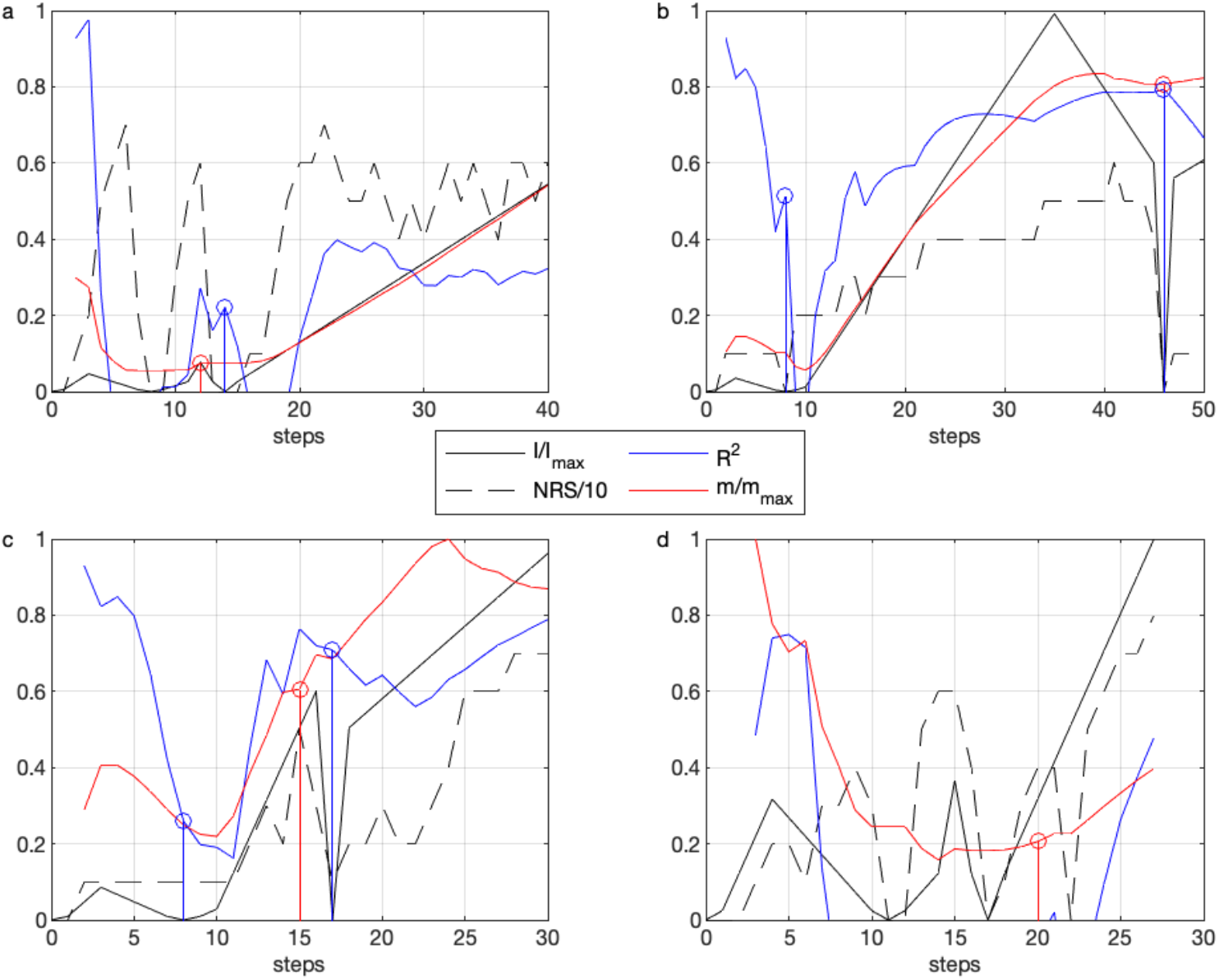
Participants rejected from further analysis. *Note*. For plot description refer to Figure 5. Rejection reasons: a. low R2 and high number of delivered stimuli; b. high stimuli current and high number of delivered stimuli; c. lack of convergence and mistake in 0mA stimulus scoring; d. lack of convergence, low R2 and mistake in 0mA stimuli scoring.

For LRM and tLRM (both methods), *R*^*2*^ values were above 0.75. No patterns could be observed in the residual plots suggesting a random distribution of residuals, implying no bias. Furthermore, the mean of residuals is close to zero and 80% of the residuals are normally distributed. The relation between stimuli intensity and NRS ratings is, therefore, linear, what is in line with our hypothesis.

Results of repeated measure ANOVA for *R*^*2*^ revealed a statistically significant main effect of the *Linear Regression Type* (F_(2,65)_ = 17.25, p<.001, *η*_*p*_ ^*2*^ =.21) and no effects of *Gender* (F_(1,64)_ =.06, p=.81, *η*_*p*_ ^*2*^ =.16) nor interaction effect *Linear Regression Type * Gender* (F_(1,64)_ = 2.10, p=.15, *η*_*p*_ ^*2*^ =.02). The pairwise comparisons of *Linear Regression Type* indicated statistically significant differences between LRM vs. tLRMm, LRM vs. tLRMmc and tLRMm vs. tLRMmc. The results indicate that the tLRM models (tLRMmc reported higher R^2^ than tLRMm, as would be expected due to a higher number of degrees of freedom) ensure the highest degree of linearity, as was hypothesized (see Table 1).

**Table 1.** Descriptive statistics of investigated variables for all four calibration models: TM, tLRMm, tLRMmc and LRM.

| Calibration method | Gender | t<sup>a</sup><br>(Mean±SD) | T<sup>b</sup><br>(Mean±SD) | Mid-painful stimulation<sup>c</sup><br>(Mean±SD) | Steps number<sup>d</sup><br>(Mean±SD) | R<sup>2e</sup> | N<sup>f</sup> |
| --- | --- | --- | --- | --- | --- | --- | --- |
| TM | Female | 0.02±0.01 | 0.26±0.32 | 0.40±0.23 | 37.68±2.64 | N/A | 31 |
|  | Male | 0.03±0.02 | 0.37±0.15 | 0.56±0.22 | 46.56±2.49 |  | 35 |
|  | Total | 0.02±0.02 | 0.32±0.16 | 0.48 ±0.24 | 42.38±1.88 |  | 66 |
| LRM | Female | 0.04±0.03 | 0.23±0.16 | 0.38±0.23 | 37.68±2.64 | 0.80±0.09 | 31 |
|  | Male | 0.07±0.05 | 0.35±0.16 | 0.56±0.22 | 46.56±2.49 | 0.75±0.15 | 35 |
|  | Total | 0.05±0.05 | 0.29±0.17 | 0.47±0.23 | 42.38±1.88 | 0.78±0.13 | 66 |
| tLRMm | Female | 0.04±0.02 | 0.19±0.10 | 0.30±0.14 | 23.55±0.75 | 0.81±0.12 | 31 |
|  | Male | 0.05±0.02 | 0.27±0.11 | 0.43±0.16 | 26.69±1.1.7 | 0.81±0.13 | 35 |
|  | Total | 0.05±0.02 | 0.23±0.11 | 0.36±0.16 | 25.21±0.73 | 0.81±0.12 | 66 |
| tLRMmc | Female | 0.04±0.02 | 0.19±0.10 | 0.30±0.14 | 23.55±0.75 | 0.83±0.11 | 31 |
|  | Male | 0.05±0.03 | 0.27±0.11 | 0.43±0.16 | 26.69±1.1.7 | 0.83±0.13 | 35 |
|  | Total | 0.04±0.03 | 0.23±0.11 | 0.37±0.16 | 25.21±0.73 | 0.83±0.12 | 66 |

<sup>a</sup> Sensation threshold
<sup>b</sup> Pain threshold
<sup>c</sup> Pain stimulation corresponding to 1.5\*T for TM and NRS of 8 for all linear models
<sup>d</sup> Number of steps/stimuli of each calibration
<sup>e</sup> Coefficient of determination
<sup>f</sup> Participant number

Wilcoxon signed rank test for the number of stimuli required by LRM vs tLRM revealed a statistically significant difference (p<0.001). tLRM take on average 17.2 steps less than LRM (36 %), what is in line with our hypothesis. Descriptive statistics of the number of steps for each calibration methods are presented in Table 1.

The results of repeated measures ANOVA for individual threshold values of *T* and *t* are presented in Table 2. We found statistically significant main effects of *Calibration Type, Threshold Type* and *Gender*, as well as interaction effects of *Calibration Type* Threshold Type* and *Threshold Type*Gender*.

**Table 2.** The results of repeated measures ANOVA for pain and sensory thresholds.

| **Main and interactions effects** | **F** | **df** | **P** | **$\eta_p^2$** |
| --- | --- | --- | --- | --- |
| *Calibration Type* | 19.06 | 3,64 | **<0.001** | 0.23 |
| *Threshold Type* | 326,74 | 1,64 | **<0.001** | 0.83 |
| *Gender* | 11.34 | 1,64 | **0.001** | 0.15 |
| *Calibration Type\*Threshold Type* | 37.42 | 3,64 | **<0.001** | 0.37 |
| *Calibration Type\*Gender* | 1.5 | 3,64 | 0.21 | 0.02 |
| *Threshold Type\*Gender* | 9.89 | 1,64 | **<0.01** | 0.13 |
| *Calibration Type\*Threshold Type\*Gender* | 1.51 | 3,64 | 0.23 | 0.02 |
| **Within-factor multiple comparison tests for T<sup>a</sup> values** |  |  |  |  |
| *Calibration Type\*Threshold Type* | **F** | **df** | **P** | **$\eta_p^2$** |
| LRM vs TM |  |  | **<0.001** |  |
| tLRMm vs TM |  |  | **<0.001** |  |
| tLRMmc vs TM |  |  | **<0.001** |  |
| tLRMm vs tLRMmc |  |  | 0.10 |  |
| tLRMm vs LRM |  |  | **0.001** |  |
| tLRMmc vs LRM |  |  | **<0.01** |  |
| **Within-factor multiple comparison tests for t<sup>b</sup> values** |  |  |  |  |
| *Calibration Type\*Threshold Type* | **F** | **df** | **P** | **$\eta_p^2$** |
| LRM vs TM |  |  | **<0.001** |  |
| tLRMm vs TM |  |  | **<0.001** |  |
| tLRMmc vs TM |  |  | **<0.001** |  |
| tLRMm vs tLRMmc |  |  | 0.42 |  |
| tLRMm vs LRM |  |  | 0.81 |  |
| tLRMmc vs LRM |  |  | 0.14 |  |
| **Between-factor multiple comparison** |  |  |  |  |
| *Threshold Type\*Gender* | **F** | **df** | **P** | **$\eta_p^2$** |
| T<sup>a</sup> female vs males | 10.90 | 1,64 | **<0.01** | 0.15 |
| t<sup>b</sup> females vs males | 8.07 | 1,64 | **<0.01** | 0.11 |

*Note.* TM: Threshold Method; tLRMm: Truncated Linear Regression Method that assumes that the coefficient *c* (the intercept) is zero; tLRMmc: Truncated Linear Regression Method where the coefficient *c* is not equal to zero;
<sup>a</sup> pain threshold
<sup>b</sup> sensation threshold

The within-factor multiple comparison tests for the interaction *Calibration Type*Threshold Type* for the *T* values showed statistically significant difference between LRM vs. TM, tLRMmc vs. TM, tLRMm vs. TM, tLRMm vs. LRM, tLRMmc vs. LRM (see Table 2 and Figure 7), what is in line with our hypothesis that *T* values calculated for longer lasting TM and LRM may be higher than ones calculated based on the tLRM. Analogous exploratory analysis performed for *t* values revealed that there was a statistically significant effect in the comparison between TM and the three types of LRM (see Tables 2 and 4 and Figure 7). It is interesting to note that for four participants the LRM method predicted negative stimuli intensity for *t*.

**Figure 7.**
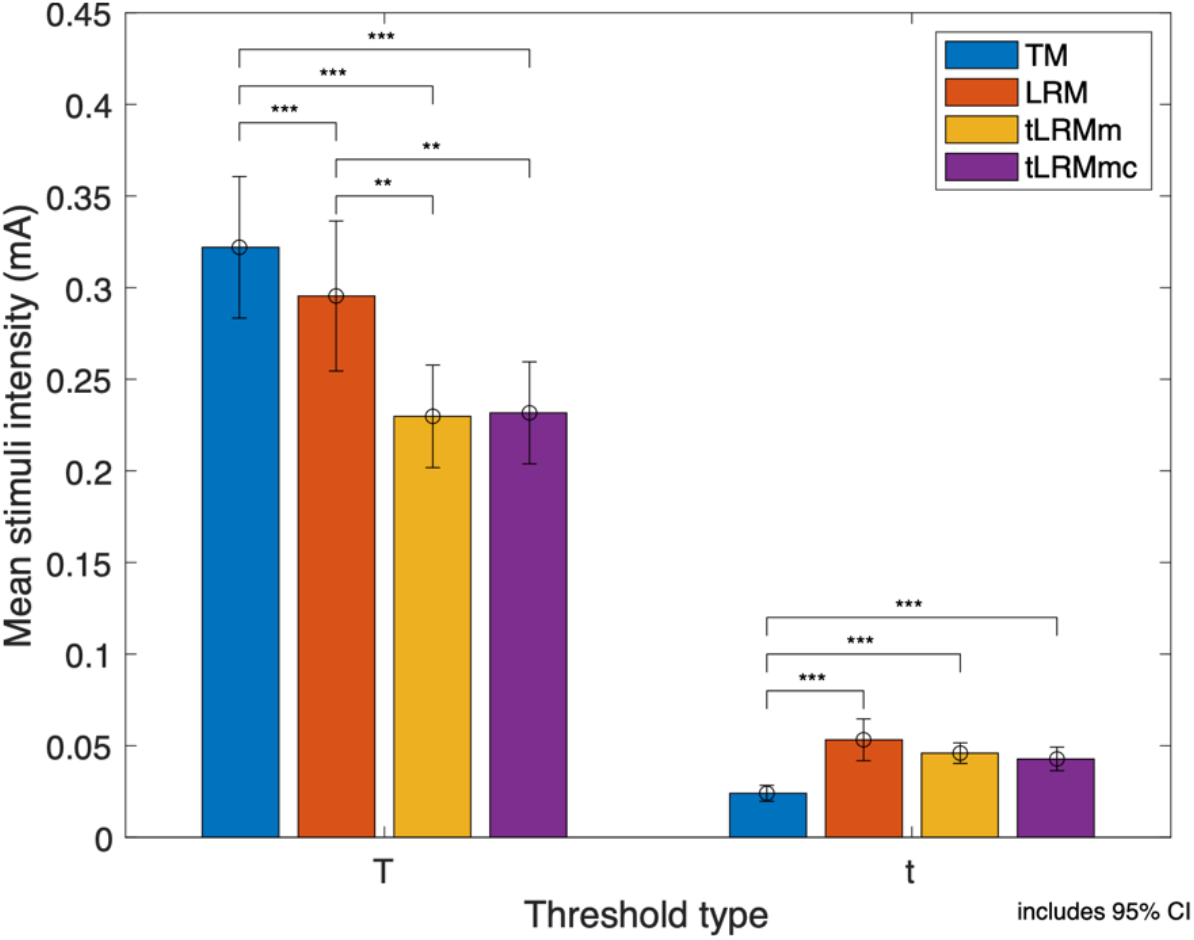
Pain (T) and sensation threshold (t) values in mA for each of the four tested calibration methods. *Note*. ** and *** corresponds to p<0.01 and p<0.001 respectively.

The pairwise comparison between the tLRMm and tLRMmc for *t* and *T* found no differences, what is in line with our hypothesis.

The within-factor multiple comparison tests showed sex differences as reflected by the interaction *Threshold Type*Gender*. For both *T* and *t*, we found statistically significant differences between both genders, what is in line with our hypothesis of the existence of gender differences in *t* and *T*. The results are presented in Table 2 and Figure 8.

**Figure 8.**
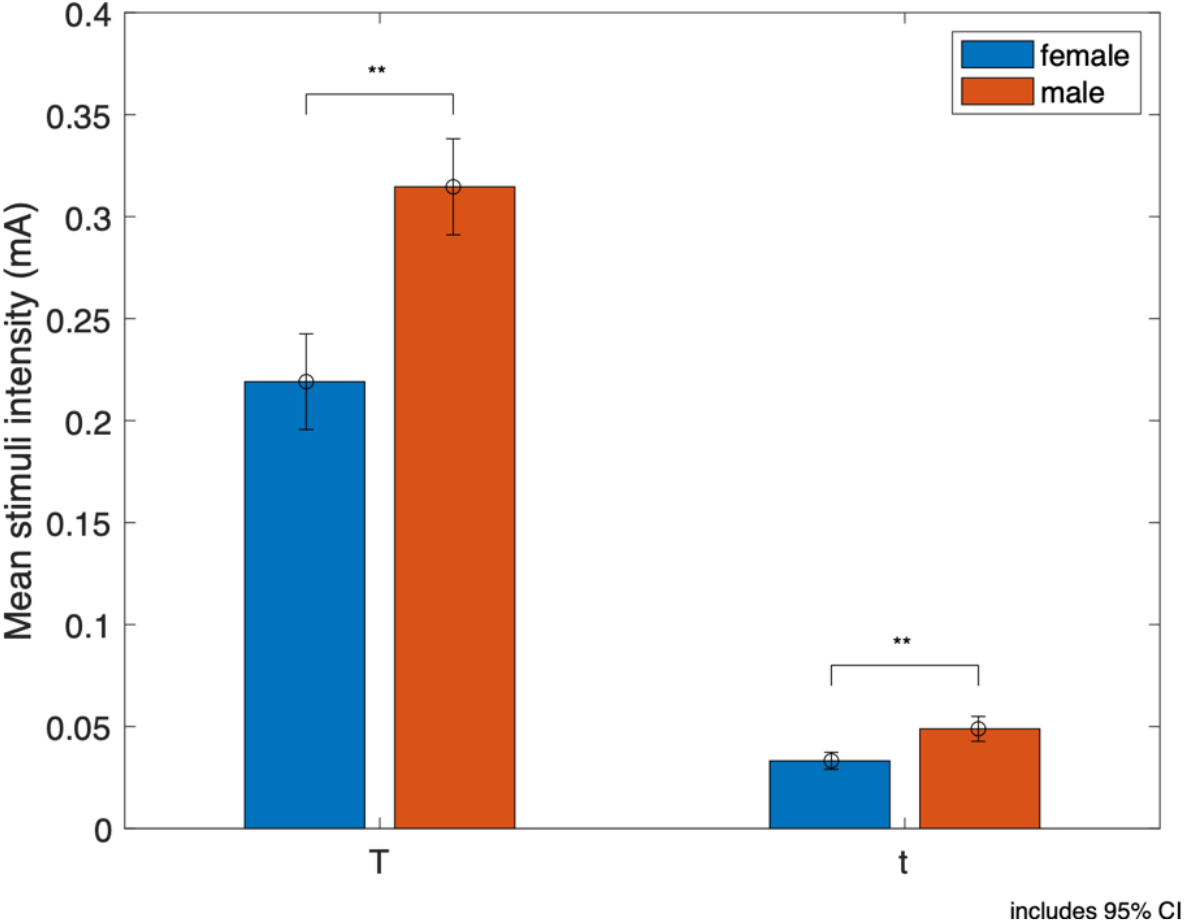
T and t values for males and females. Note. ** corresponds to p<0.01

The ANOVA analysis for mid-painful stimulation indicated statistically significant main effects of *Calibration Type* (F_(3,64)_ =19.38, p<.001, *η*_*p*_ ^*2*^ =.23) and *Gender* (F_(1,64)_ = 10.63, p>.01, *η*_*p*_ ^*2*^ =.14), but no interaction effect *Calibration Type* Gender* (F_(1,64)_ =0.93, p> .05, *η*_*p*_ ^*2*^ =.01) (see Figure 9 and Table 1).

**Figure 9.**
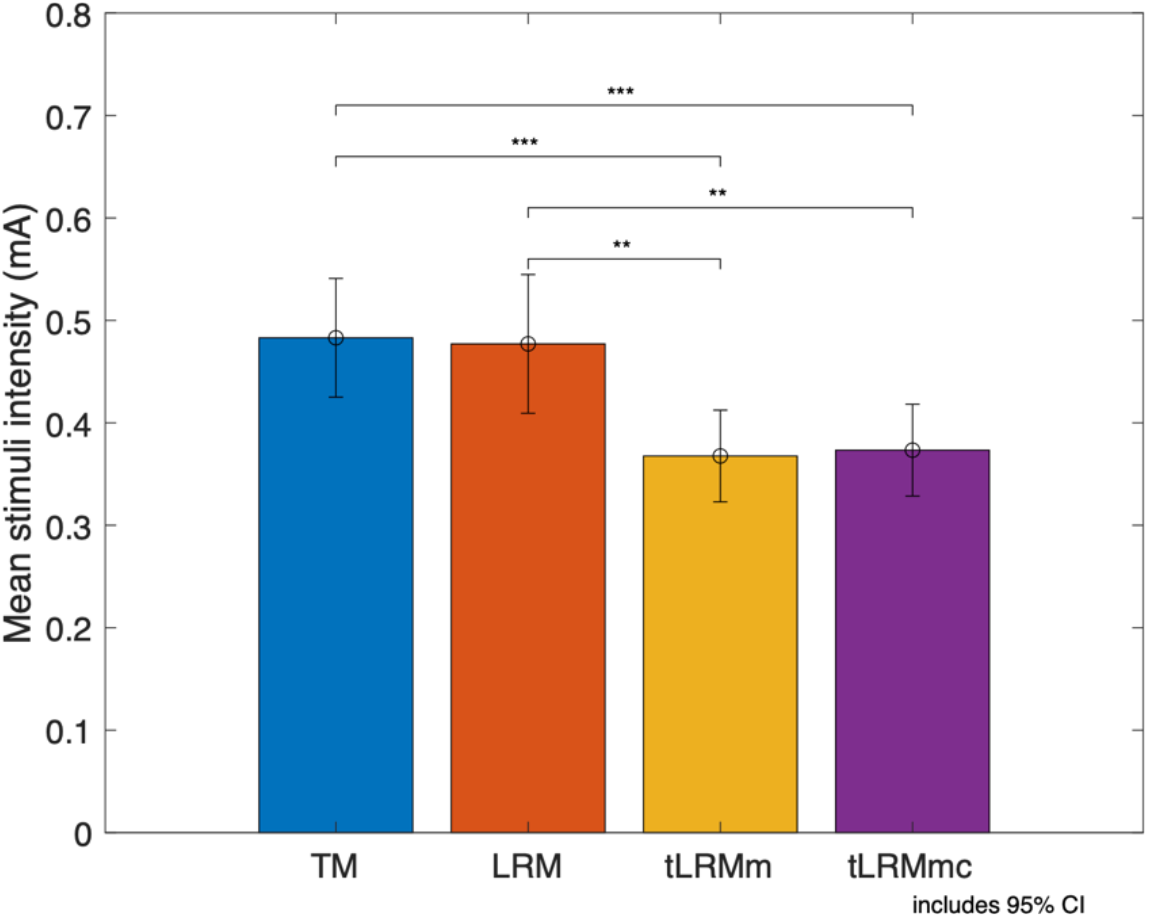
Mid-painful stimulation estimated for each of four calibration methods. *Note*. For TM the equation 1.5T was used to set mid-painful stimulation. ** and *** corresponds to p<0.01 and p<0.001 respectively. For TM the equation 1.5T was used to set mid-painful stimulation.

The pairwise comparisons indicate a statistically significant difference (p<.05) between TH vs. LRM, TH vs. LRMm, TH vs. LRMmc, LRM vs. LRMm, and LRM vs. LRMmc. These results are in line with our hypothesis that TM and LRM may overestimate the intensity of the pain stimuli.

The post-test revealed that, for most of the participants, mid-painful stimulation was indicated for NRS scores between 6-9, low-painful between 3-6, and tactile between 2-3.

## 5. Discussion and conclusions

Our aim was to develop an automatic calibration method that takes advantage of LRM and reduces the number of delivered stimulation. We highlight the importance of calibration data quality check, compare different calibration methods, and offer complete laboratory set up that can serve further investigators. Our extensive literature review has revealed no similar studies, where the methodology and laboratory set up would be provided in such detail. It is an enabling technique that serves to perform precise and reliable neurophysiological studies where the use of complex equipment, such as MEG and/or EEG in conjunction with WASP electrodes, is necessary.

In line with our hypothesis, the relationship between NRS scores and stimulus intensity was shown to be highly linear, which is indicated by high *R*^*2*^ values for LRM and both tLRM models, confirming the results of previous studies^12,13^. Linearity of the stimuli intensity and its ratings is under the debate for a long time and lack of consistency in previous studies^14,15^ and our result may be associated with the investigated laboratory stimulation type and the specific clinical pain type^17^.

Furthermore, tLRM shows higher *R*^*2*^ values, indicating that considering only an optimal dataset gives better results than a complete one, likely due to habituation effects in the latter. In the study we used a 11-point NRS where both non-painful and painful stimuli were rated^20,21,34^ which, together with tLRM, allows for performing estimations on stimuli intensity and NRS ratings. Finally, truncating data has another advantage, namely decreasing the number of delivered stimuli. This means that the tLRM required on average 17.2 less stimuli making it less painful and less uncomfortable for the participant, and possibly mitigating habituation.

Since data linearity has been shown, *R*^*2*^ can be used to quantify the correctness and reliability of the calibration procedure. This allows for an automatic (no human labour required) and instant (immediately after calibration end) procedure for participants rejection according to the three proposed criteria. As a result, the experimenter can avoid perpetuating errors in further stages of the experiment, which can save time and financial resources.

Further reliability investigations involved threshold comparisons between the methods. On the one hand, LRM and tLRM yield statistically similar results for *t*, which are higher than for the TM method. This means that either the linear regression models overestimate the required stimulus, or the TM underestimates it. We suspect that the latter is more probable, due to a higher sensitivity to stimuli at the beginning of the procedure when *t* is predicted by TM. On the other hand, only tLRMm and tLMRmc yield statistically similar results for *T*, but their mean is lower than for TM and LRM. This is in line with our hypothesis that, in general, TM and LRM would indicate higher stimuli intensity as they require more stimuli application and therefore participants can suffer from habituation. Habituation can be minimized by using an adequate ISI, which in our study was at least 8 sec. In studies using analogous electrodes (IES) the rest time between three stimulation sets was 1 min^37^ or an ISI between 2.5 and 3.5 sec^38^. Consequently, the chosen ISI was sufficient, but in future studies including breaks between ascending/descending curves can further mitigate those effects. Future verification method could also include applying an extra set of random stimuli and comparing their ratings with the predictions.

Next, since tLRMm and tLRMmc yield statistically similar results for *t, T*, and mid-painful stimuli, we propose to use the tLRMm method, as it is described by only one parameter, *m*, and forces the condition that for no stimulation the NRS is 0. This, in turn, makes comparisons between participants and experiments easier, and avoids not realistic situations where negative stimulation is predicted for low NRS, as was observed for a few cases of LRM. Regarding gender differences, our results indicate that females require a lower stimulation intensity than men to reach both the sensation and painful thresholds. This may suggest that women have a higher sensory acuity, at least in response to electric stimulation via WASP electrodes. This conclusion is not consistent across other studies^39,40^ ; however, the predominant view is that women have a greater pain responsiveness for most pain modalities^41^. This is not unique to pain, as there is evidence that women are often more perceptive across multiple sensory domains^23^, exhibiting a greater detection and discrimination sensitivity to tactile^42^, olfactory^43^, and visual stimuli^44^. Our results are also in line with the clinical reality which has shown that gender differences exists with respect to pain tolerance and thresholds, and that there is a higher prevalence of chronic pain conditions for females^41,45,46^. Finally, gender differences in pain perception and modulation exists at molecular, cellular, and system level^47^.

In relation to the above, the calibration procedure was made as automatic as possible in order to improve standardization and avoid further experimenter biases^48^. Moreover, to control the effect of experimenter characteristics and gender, a woman and a man were always present in the laboratory. The literature shows significant ambiguity in that topic, and it is possible that pain modality may play a crucial role. For instance, previous studies indicate higher thresholds for electric pain determined in the presence of a female investigator regarding participant gender^48^. Another study, where pressure stimuli was used, shows that men showed higher average pain thresholds when tested by a female experimenter^49^. Interestingly, heat pain thresholds were not shown to be significantly influenced by experimenter gender^50^. For this reason, in future studies it is recommended to investigate the role of experimenter gender on pain thresholds produced via WASP electrodes.

The last analysis aimed to investigate the assumption that non-painful stimuli to make predictions about painful stimuli and it involved a comparison between predicted levels for mid-painful stimulus. There is no statistically significant difference between TM based on 1.5\**T* and LRM. However, they are different to the intensity levels predicted by tLRM, which are significantly lower, possibly due to habituation. However, we cannot exclude that the tLRM mid-painful intensity is underestimated, as the post test for LRM validated its correctness with low- and mid-painful stimulations. A possible way of mitigating this effect could be using an adaptive staircase method^16^. This method first applies several stimuli, which serves to create the initial linear model, followed by a set of stimuli with random order and strength, aimed to refine that model. Such a refinement, based on random stimuli, could be added to the tLRM method to verify the possibility of underestimation of the threshold-related stimuli. Nevertheless, the higher degree of linearity in tLRM is a strong indication that non-painful stimulation calibration can be extrapolated to provide predictions of painful stimulation. However, to further improve the proposed calibration reliability, a random strength stimulation verification of reliability should be performed *a posteriori*. This could be used to check if the calibration results are consistent over time, as was done before for thermal stimulation^36^.

Concluding, the proposed method, including its correctness quantification and rejection criteria, can be easily adapted to other stimulation methods, and to any neurophysiological studies on pain. We established procedures for the application of the stimulus as well as a specific automatic calibration of electric stimulation delivered via WASP electrodes. Additionally, we provide Matlab and LabView scripts that can serve further studies. We believe that the replicability of the laboratory setup is crucial for further comparison of study results and development of reliable psychophysiological pain studies. The importance of proposed calibration encompasses also clinical studies where experimental pain models are used, for instance, to test the effectiveness of analgesic compounds^51^. Furthermore, according to multi-modal approach in human pain research^52^, this method also allows for a standardized comparison between different stimulation modalities, facilitating translation of their result to clinical setting. Furthermore, linear regression satisfies the definition of the IASP, which recommends that pain threshold should be at the level at which 50% of stimuli would be recognized as painful instead of the least stimulus intensity at which pain is perceived^53^.

Finally, our method is not free from limitations and the need for further refinements. Reliability tests are required for future experimental studies on WASP electrodes and studies where pain biomarkers or signatures are evaluated^54,55^. Furthermore, the intra-epidermal electrodes, such as WASP, have been found to be sensitive to their positioning relative to the location of the nerve fibre^38^, which could have influenced the calibration. Finally, more research is needed to determine the factors influencing the sex-based differences in *t* and *T*, such as possible differences of electrode attachment, or gender physiological, chemical, and biophysical skin differences^56^. In future studies it is suggested to use Visual Analogue Scale (VAS) that is commonly used in clinical practice^57^ and often described as a ratio scale^14,17^. However, there is data indicating a high corelation between both VAS and NRS^58^ and suggesting that the choice between the mentioned scales for pain assessment can be based on subjective preferences^59^.

## Data Availability

All data produced in the present study are available upon reasonable request to the authors

## 6. Acknowledgements

We would like to thank all the participants and Gabriel Delojo Pradas for his enthusiasm and practical support in conducting the study.

## Consent to participate

All subjects signed informed consent to participate in the study.

## Availability of data and materials

The datasets analysed during the current study and the code used during the experiment are available from the corresponding author upon request.

## Authors’ contributions

Conceptualization: KS, RB

Data curation: KS

Formal analysis: KS

Funding acquisition: KS

Investigation: KS

Mythology: KS, RB

Project administration: KS

Resources: KS

Software: KS, RB

Supervision: SM, RB

Validation: not applicable.

Visualization: KS

Writing-original draft: KS

Writing-review and editing: all authors

## Supplement materials

**Table S1.** Input signal and its corresponding action performed by the LabVIEW script

| Input | Signal duration | Action |
| --- | --- | --- |
| **Signal coming from ai0 channel only** | 0,5 sec | Increase the output voltage by 1mV (equivalent to increasing stimulation current by 0.01 mA) |
|  | 1,0 sec | Increase the output voltage by 2mV (equivalent to increasing stimulation current by 0.02 mA) |
|  | 1,5 sec | Increase the output voltage by 5mV (equivalent to increasing stimulation current by 0.05 mA) |
| **Signal coming from ai1 channel only** | 0,5 sec | Decrease the output voltage by 1mV (equivalent to decreasing stimulation current by 0.01 mA) |
|  | 1,0 sec | Decrease the output voltage by 2mV (equivalent to decreasing stimulation current by 0.02 mA) |
|  | 1,5 sec | Decrease the output voltage by 5mV (equivalent to decreasing stimulation current by 0.05 mA) |
| **Signal coming simultaneously from both ai0 and ai1 channels** | 0,3 sec | Send electrical stimulation of the previously selected intensity and 300 ms of duration. |

<sup>f</sup>Participant number

**Figure S1.**
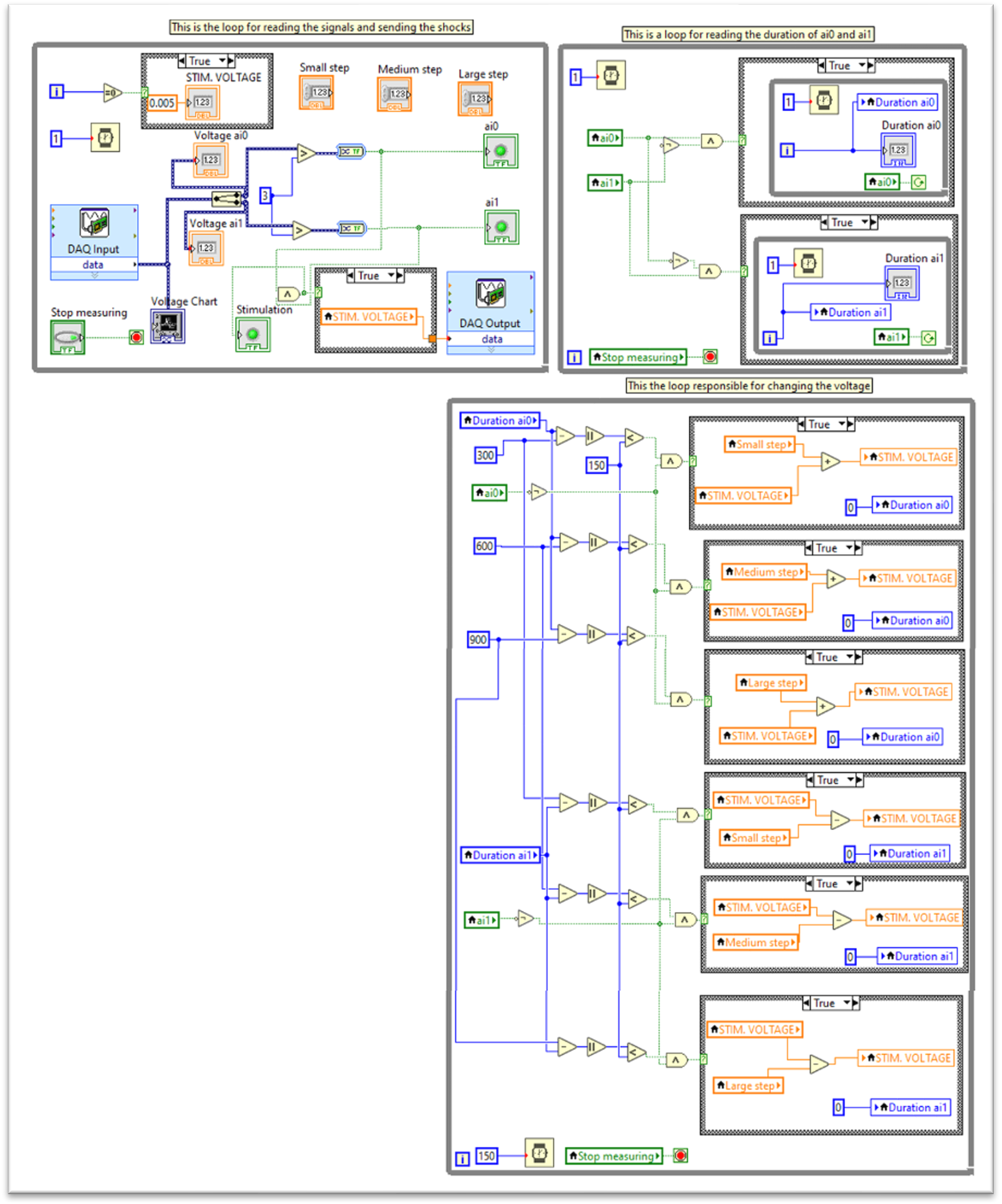
Calibration LabView design.

